# Cohort Profile: Swiss BioRef: The building blocks of a nationwide IT infrastructure in Switzerland for generating precise reference intervals

**DOI:** 10.1101/2022.08.30.22279391

**Authors:** Tobias U. Blatter, Harald Witte, Jules Fasquelle-Lopez, Jean Louis Raisaro, Alexander B. Leichtle

**Affiliations:** University Institute of Clinical Chemistry Inselspital - Bern University Hospital and University of Bern; Graduate School for Health Sciences (GHS) - University of Bern; University Hospital Lausanne (CHUV) - University of Lausanne; Center of Artificial Intelligence in Medicine (CAIM) - University of Bern

## Abstract

**Purpose:** Swiss BioRef is a nation-wide multicentre infrastructure project, the aim of which is to become a sustainable framework for the estimation and assessment of patient-group-specific reference intervals in laboratory medicine and beyond. In this unprecedented effort, nation-wide multidimensional data from multiple clinical laboratory databases has been combined under the common interoperable semantic framework of the Swiss Personalized Health Network (SPHN) initiative. The consolidated effort enables creating extremely detailed patient group-specific queries via intuitive web applications, allowing the generation of individualised, covariate adjusted reference intervals on-the-fly.

**Participants:** The project is a collaborative effort of four major hospitals in Switzerland, the University Hospital Bern (Inselspital, “Insel”), University Hospital Lausanne (CHUV), Swiss Spinal Cord Injury Cohort (“SwiSCI”) and the University Children’s Hospital Zurich (“KiSpi”), and two academic groups in Bern and in Lausanne.

**Findings to date:** Within the infrastructure we deployed, the laboratory data from four major hospitals (approximately 9 million measurements from 250’000 patients) is made available to two conceptually different web applications (one centralised and statistically detailed, one decentralised using distributed computing). They enable the inference of reference intervals for more than 40 blood test variables from clinical chemistry, haematology, point-of-care-testing, and coagulation testing, with various patient factors (such as age, sex and a combination of ICD-10 defined diagnoses) and analytical factors (such as type or unique identifiers) that can be used to generate precise reference intervals for the respective groups.

**Future plans:** Now that all required basic infrastructure elements for Swiss BioRef are deployed, we are evaluating inter-cohort transferability of semantic standards, “change tracking” in merged databases and biological variation of the blood test variables, in order to generate precise reference intervals. While adjusting the developed web-interfaces to suit the needs of the various end-users, we additionally plan to onboard new national and international partners.

**Strengths and limitations of this study:**

- The Swiss BioRef project is the first multi-cohort infrastructure in Switzerland for the estimation of precise reference intervals in laboratory medicine.
- With the BioRef consortium agreement a common framework for multi-cohort data sharing, hosting, and accessing has been thoroughly defined.
- The definition of interoperable data formats and data encoding for Swiss BioRef permits the fusion of the various data sources into a unified infrastructure. Due to differing data management systems at the individual clinical data warehouses, the harmonisation of data contributions requires significant effort which limits direct data provision.
- Two different web applications with varying data access architectures enable researchers to map the individual complexity of their patients into a substantiated statistical analysis to infer precise and highly relevant reference intervals. Needless to say, anticipating the requirements of an increasingly diverse user base remains a challenging task.
- Due to the modular expandable architecture of Swiss BioRef, potential national and international partners can easily access and even join the network.

## Introduction

The use of blood tests is a cornerstone of non-invasive disease diagnosis and health assessment in clinical medicine. When clinicians are trying to discern patients from healthy persons, they rely on population-based measures such as the reference interval (RI). In its core concept, RIs enclose a fixed range of values from a predefined reference population (e.g. 95%) and it has been long established that they are as effective in clinical use as long as they are precise and accurate[1–4]. Clinical laboratories need to independently establish and periodically verify their RIs in use by admissible guidelines[5]. The most-cited guideline, the *EP28-A3c* by the Clinical and Laboratory Standards Institute (CLSI) and the International Federation of Clinical Chemistry (IFCC) states that RIs should be estimated from cohort-relevant reference populations, where patient-group-specific covariates such as age, biological sex, ethnicity, and region are considered but also differences in pre-analytical factors are accounted for[6]. The process is cumbersome, costly, and often beyond the scope and possibilities of many independently operating laboratories: Cohort-specific analyses require stratification by a specific combination of the above-mentioned covariates. Therefore, these analyses frequently suffer from small sample sizes due to a lack of available data[7,8]. Limitations can be overcome with multicentre collaborative reference interval studies, where standardised protocols help to derive *harmonised RIs* on a national level by pooling the appropriate number of subjects from multiple cohorts[9]. Such standardisation requires clear classification systems, e.g. for the nomenclature, terminology, units and formats used, to ensure reproducibility of all the steps of the total laboratory testing procedure, possibly for international application[10,11]. This is an on-going global process, as laboratories in Europe[12–17], Africa[18–20], North America[21], Asia[22–24] and Australia[25] aim to derive nation-specific RIs with multicentre studies.

Despite this welcome effort, most studies have been conducted with very lenient inclusion/exclusion criteria due to a missing overarching definition of “health”, covering both the normative aspects (well-being and functioning) and more descriptive aspects of health evaluation (test result assessment). This hinders the comparability of generated RIs[26]. A common classification framework to define the health status of included subjects based on predetermined medical conditions is needed. The International Classification of Diseases (ICD) coding system is one coding system to help represent nuanced diseases to broader morbidities[27]. We suggest that being able to map individual combinations of multiple morbidities of patients on the local analysis of RIs will help to establish more formalised RIs for precision medicine. Even so, the broader introduction of locally inferred RIs has not been seen across the board in clinical laboratories, mostly due to a lack of sophisticated analysis tools connecting laboratory databases, where multi-dimensional data is readily available, to physicians in need for clinically relevant RIs. The *Swiss BioRef* rationale is to establish an inter-cohort infrastructure, to create precise RIs from pooled data based on an interoperable semantic framework. Establishing an opportunity for clinical laboratories to check whether their standard RIs apply to their patient populations in a convenient and reproducible way, should be a vital part of practised precision medicine today.

## Cohort Description

### Swiss BioRef project setup

The Swiss BioRef project is an ongoing multi-cohort project, which aims at creating the necessary computational resources to infer precise RIs on-the-fly based on multiple covariates for the application in clinical medicine. The initial grant proposal for the Swiss BioRef project was peer-review approved in 2018 by the Swiss Personalized Health Network (SPHN) as an infrastructure development project, falling under the scope of the development and testing of new infrastructures for personalised health related research[28]. Swiss BioRef requires a secure IT network infrastructure, where clinical laboratory data from various clinical data providers is linked to easy-accessible web-interfaces which allow the recurrent aggregation of patient data from all partners in an accreditation-proof way. Efforts towards inter-cohort alignment of sensitive health-related data have been promoted by the Data Coordination Center (DCC), a core part of SPHN. Along those lines, a national IT-environment for sensitive research data, the BioMedIT infrastructure, has been established to ensure a backbone for secure transfer, storage, management, and processing of confidential data[29]. The BioMedIT infrastructure is a collaborative effort of three Swiss partners, the Core□IT at the Swiss Institute of Bioinformatics (SIB) in Lausanne, the Scientific IT Services (SIS) at ETH Zürich, and the centre for scientific computing (sciCORE) at the University of Basel, managed by the DCC. Data transfers within this network follow the “snowflake” principle: One BioMedIT node serves as the main or destination node whereas the other nodes act as relays or transfer nodes. Data providers simply send data to their closest node from where it is routed to the destination node (in case of transfer node)[30]. For Swiss BioRef, the BioMedIT node at the ETH Zürich serves as the main node.

Based on common laboratory semantics and under a common contractual architecture, the Swiss BioRef project includes two different mechanisms for the multi-cohort data pooling, a centralised approach and a decentralised approach. Aside from the chosen approach, it is vital that each clinical partner involved is willing to process the data to adhere to the harmonised and interoperable standards for the data encoding. Overall, the Swiss BioRef project enables insights into the challenges of a multi-centre approach for RI inference, i.e. the inter-cohort transferability of semantic standards, “change tracking” in merged databases and biological variation of the blood test variables.

### BioRef consortium

The parties involved in Swiss BioRef decided to join efforts in the form of a multicentre research consortium - the BioRef consortium - to oversee the project. As of August 2022, the BioRef consortium contains the University Hospital in Bern, («Inselspital», Bern), the University Children’s Hospital in Zurich («KiSpi», Zurich), the Swiss Paraplegic Research («SPF», Nottwil) and the University Hospital in Lausanne (Centre Hospitalier Universitaire Vaudois, «CHUV», Lausanne). The consortium agreement (CA) covers the data governance, data delivery and required network infrastructure as well as the division of labour regarding the development of the web interfaces to infer RIs from Swiss BioRef data. Substantial support to establish the CA, tailored to the requirements of the Swiss BioRef project, was provided by the SIB and DCC. The Swiss BioRef project was led by the computational medicine group in Bern (<u>Computational Medicine Group</u>) which developed the necessary IT-components in collaboration with the health informatics and data privacy group in Lausanne (<u>Health Informatics and Data Privacy</u>). These components were deployed with the help of the SIS at ETH Zürich representing the BioMedIT node in Zurich. The CA includes additional legal compliance agreements, a data transfer and use agreement (DTUA), and a data transfer and processing agreement (DTPA), stating the three BioMedIT nodes in Zurich, Basel, and Lausanne as parties responsible for transfer, hosting, and processing of the data. Services required to set up the Swiss BioRef architecture were rather complex, therefore an additional service level agreement was concluded with the SIS to cover effort exceeding the base services of BioMedIT. Last but not least, the CA also covers the process for future partners to join, making the overall consortium architecture expandable in a modular way, even beyond national borders, while still adhering to the defined rules and permissions of the multi-cohort data sharing project.

### Data governance and regulatory aspects

Laboratory data is considered “sensitive data” and requires careful governance, covered by the Swiss Federal Act on Data Protection (FADP 1992, art. 3c) and the Human Research Act (HFG RS 810.30). Research conducted with non-anonymized laboratory data requires the approval from the cantonal ethics committee (CEC). For the use of anonymized data, however, the HFG does not apply. Therefore, studies respecting patient consent and data protection provisions as specified in the FADP do not require approval from the CEC. For inter-cohort data sharing a Data Transfer and Use Agreement (DTUA), in line with the FADP and HFG, must be concluded between the data provider and the recipient before any sensitive data can be exchanged. The hurdle for nation-wide data pooling is therefore relatively high. A recent effort of establishing a Swiss multi-cohort resource in pharmacogenetics has been documented to last up to a full year for only setting up the legal and scientific framework[31]. The CA defines that each party hosts their respective laboratory data provided for the project either on the secure IT infrastructure BioMedIT network in a dedicated project space on Leonhard Med (LeoMed) for the purposes of the project or locally by using MedCo, a secure system for privacy-preserving federated analytics based on multiparty homomorphic encryption, where the data stays locally at the particular site of the participating institution and only the aggregated result of the requested computation is released in cleartext to the authorised user[32]. All parties must process the data in compliance with applicable data protection laws, including consent regulations. The CA expresses that each participating partner only has unrestricted access to their own data. Access to data originating from multiple cohorts, however, is permitted in the form of insensitive data aggregates. These are sufficient for the estimation of RIs and are generated according to the cohort queries of the user. Any other access to Swiss BioRef data is regulated by the BioRef CA. This precludes access to raw data for external researchers and special conditions for accessing the analysis tools working (GUI web applications) apply.

### Data interoperability

The key component for creating a sustainable and expandable infrastructure is the definition of inter-cohort concepts regarding the interoperability, the availability, and the dimensionality and quality of the data provided by different cohorts. In an impressive effort, the use of LOINC (Logical Observation Identifiers Names and Codes (LOINC)[33]) as standard in encoding laboratory data has been achieved by the SPHN-funded L4CHLAB project across the Swiss university hospitals. About 1’500 laboratory analyses have been mapped to LOINC coding [34,35], improving clarity of encoding and enriching results with valuable meta-data. If all these conditions regarding interoperability and privacy are met, reference populations can be pooled from multiple cohort databases when establishing RIs.

Consortium partners extract their data set on their own accord, giving them full control over the data sharing process. They are only urged to adhere to the predefined and agreed upon LOINC encoding. As a preferred data transfer logic, the Resource Description Framework (RDF) was chosen. In collaboration with the DCC, the Swiss BioRef ontology was established based on the SPHN ontology to ensure correct conversion of data to RDF-format[36]. Participating sites could also contribute their data in a csv (comma separated value) - format if they include the aforementioned metadata to each data point.

### Measurements

Data from each contributing cohort consists of laboratory test results («measurements») from more than 40 laboratory variables, uniquely defined by LOINC encoding. The following metadata is currently included with each measurement: Patient record information such as the age (in years), the administrative sex, the five most relevant previously established diagnoses using the ICD-10-GM-encoding[27], and information on the generation of the measurement (pre-analytical factors). The latter specifies the analyser and the used test kit/reagent by the unique identifiers for medical devices from the Global Unique Device Identification Database (GUDID)[37] as well as the type identifiers from the Global Medical Device Nomenclature (GMDN)[38]. This additional metadata shall help to overcome the lack or sparsity of information LOINCs offer with respect to the applied method.

### Data delivery

The Swiss BioRef project pursues two separate mechanisms for inter-cohort data sharing. The first mechanism is the privacy-preserving system MedCo, an operational system which uses a multiparty homomorphic encryption scheme and obfuscation techniques. MedCo relies on a decentralised peer-to-peer infrastructure, enabling the processing of sensitive data under encryption and the release of results aggregated across all participating sites[32]. MedCo follows a strict “no copy, no move” principle, where no clinical data must leave the local site’s database and only aggregates are exchanged and further processed between different nodes, always under encryption. The second mechanism for inter-cohort data sharing - allowing for more tailored analyses - relies on the secure BioMedIT network, where data from BioRef consortium partners are separately collected in a highly restricted central database. This data transfer is coordinated by the SPHN Data Coordination Centre (DCC), the central office for the BioMedIT network. Data providers use the Secure Encryption and Transfer Tool (SETT) of BioMedIT[39], which encrypts the data, signs this process, and carries out the transfer to the BiomedIT nodes. The cryptographic keys of the data provider and data manager of the project are stored on a specific key server of the DCC. The encryption guarantees minimal exposure of sensitive information during data transfer.

## Findings to date

### Deployed infrastructure

The requirements, such as the setting up the CA, harmonising the data to ensure interoperability and defining the data delivery processes, enabled the setup of a comprehensive and stream-lined process to request patient queries from multiple clinical cohorts. In the following, we present the Swiss BioRef infrastructure, currently deployed with data contributions from four major Swiss hospitals and embedded in the nationwide secure environment of the BioMedIT computing network. Our infrastructure allows the aggregation of clinical data in a unified manner to create a comprehensive database resource (Figure 1).

**Figure 1:**
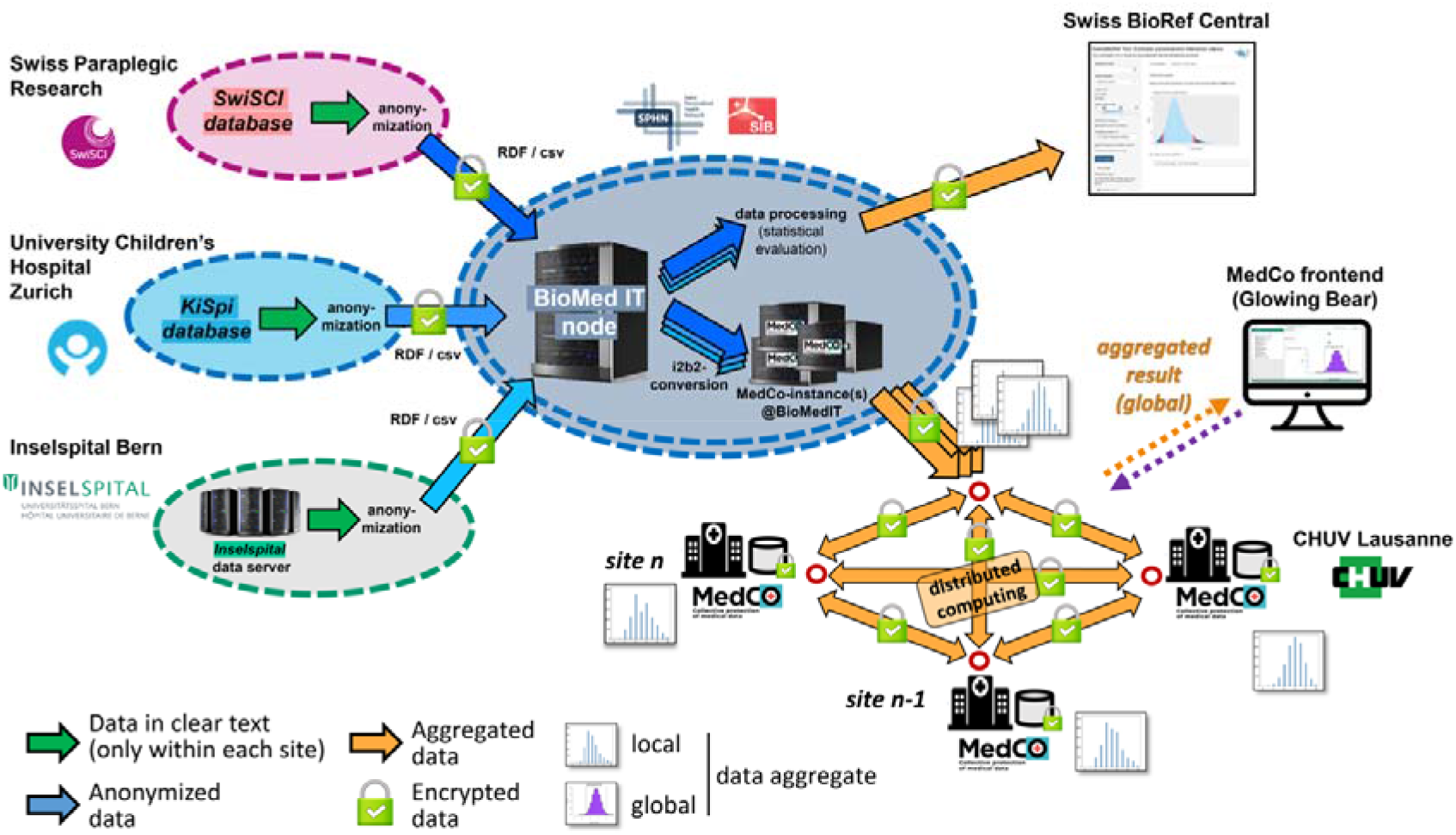
Illustrative representation of the *Swiss BioRef* infrastructure: Data is either transferred from individual data providers, aggregated, and analysed within the BioMedIT infrastructure. Alternatively, data providers using the MedCo access route (depicted by the distributed computing network) host their data on their own premises and share encrypted aggregate data only. Data processing and query computing to present reference intervals (RIs) is then carried out in two separate user-facing web applications, “Swiss BioRef Central” and “Swiss BioRef - MedCo”.

The first part of the deployed infrastructure relies on the secure and encrypted data retrieval using the established BioMedIT network. Before data transfer, the data providers from SPF, KiSpi, and Insel were requested to anonymize and provide their data in the agreed upon RDF format or a suitable tabular format such as the csv-format. After a rigorous quality control, the anonymized data set was transferred between the participating institutions and the BioMed node in Zurich by using SETT[39]. The transfer, hosting, and processing of data from the SPF, the KiSpi and Insel were all carried out in the secure network of BioMedIT and retrieved in the dedicated project space on LeoMed at the BioMedIT node in Zurich. Upon data retrieval at the BioMed node in Zurich, the data were decrypted by the Swiss BioRef data manager. The pooled standardised data creates the multi-cohort database readily available within the Swiss BioRef project space. The data of CHUV was made available directly by the MedCo system. The transfers were initiated after the signing of the CA, which covered the DTUA and the DTPA. The database was then pre-processed by the data manager for the Swiss BioRef Central web application and for the MedCo system architecture.

### Web interfaces

To compare a decentralised and centralised approach for the analysis of multi-cohort data, two separate web applications with varying system architectures have been set up.

The first deployment architecture is built on the MedCo operational system (“Swiss BioRef MedCo”, Figure 1, p. 8). MedCo, developed by researchers and engineers from the École Polytechnique Fédérale de Lausanne (EPFL) and the CHUV, offers a decentralised way of analysing medical data from multiple cohorts[32,40]. The use of the MedCo architecture circumvents the necessity to store data in a single central repository. By using homomorphic encryption, it allows the computation on encrypted data aggregates queried from multiple separate participating cohorts, enabling trust decentralisation and end-to-end confidentiality protection. Patient-level data stays on site at each participating institution, regardless of where these are located. With this, the MedCo system forms a secure, federated and interoperable network that is both compliant with current legislation of handling of sensitive patient data, as no personal data is ever shared, and fulfils the same interoperability standards set out in the aforementioned centralised approach of medical data sharing within the BioMedIT infrastructure[32]. The extended MedCo system in context of the Swiss BioRef project contains the MedCo back end and a modified MedCo front-end. On the back end, MedCo is built in a separate but modular approach, in which the query-system never accesses the unencrypted data stored in the i2b2 common data model (Informatics for Integrating Biology and the Bedside), implemented in over 300 hospital cohorts worldwide[41]. This mechanism enables full end-to-end data protection. A data converter module automatically translates the data from RDF to i2b2-format[42]. A connector module then processes incoming or outgoing queries. Lastly, a cryptographic module wraps incoming and outgoing queries with the necessary encryption. The encryption protocols used in MedCo are based on the UnLynx crypto library[43]. The front-end of the MedCo system is the user-facing web application based on Glowing Bear, a GUI for clinical data selection and analysis[44]. MedCo has adapted its Glowing Bear interface to adjust the i2b2 framework and enable distributed queries and the homomorphic encryption and decryption capabilities. For Swiss BioRef, the Glowing Bear interface has been tailored further to allow the generation and visualisation of precise RIs using an IFCC/CLSI approved method for non-parametric RI estimation.

The second deployment uses a containerized approach to run an R Shiny application within LeoMed (“Swiss BioRef Central”, Figure 1, p. 8). R Shiny is an operative extension of the R programming language into web application development to allow reactive and interactive data analysis[45]. In the overall structure, the R Shiny web application serves as a development and validation tool for the Swiss BioRef team to test the performance and accuracy of the statistical methods on inferring precise RIs from multi-cohort resources. The application has both direct methods (IFCC/CLSI approved[6]) and indirect methods (using newer data mining techniques[46]) for the inference of RIs implemented. The web application is running on a virtual machine (VM) within the LeoMed network with full access to the anonymized data stored as .csv files. The GUI of the application interactively allows for setting and executing patient queries based on the covariates defined by the BioRef consortium client-side and running an appropriate statistical inference method on the returned measurements server-side. Only the calculated RIs with a bootstrapped 90% confidence interval and pre-build histograms are rendered to the GUI for the end-user, preventing malicious data breaches of the data set loaded in the back end. The web traffic of Swiss BioRef Central is implemented behind a reverse-proxy layer in the application architecture. This hides server traffic and communication to the front-end of the application, which further reduces the risk of exposing sensitive information to the front-end.

### Explicit data governance

The CA defines that the access to the web applications is granted by the executive board of the BioRef consortium. End users are only given access to visual aggregates of the original data and the RIs. To streamline the user-management, a separate Swiss BioRef end-user licence agreement (EULA), which defines the terms and conditions that apply for clinical end-users to access the deployed web applications, was defined. As of today, researchers and clinicians with a valid SWITCH edu-ID account can apply for registration. The SWITCH edu-ID is part of the Swiss SWITCH authentication and authorisation infrastructure (SWITCHaai), where researchers are given a lifelong and user-centric identity based on their association with a university[47]. This simplifies the user management process as verification of new users can be easily automated.

## Future perspectives

The evaluation of the two web applications within the LeoMed environment is ongoing in terms of performance and accuracy of the statistical analyses employed. A report on the first harmonised RIs from a multicohort approach in Switzerland is currently in preparation.

### Expandability

With the establishment of the BioRef consortium, an interoperable and secure framework to pool multidimensional laboratory data from various cohorts into a valuable database resource of laboratory measurements in Switzerland has been created. As the Swiss BioRef framework can be expanded in a modular way, the inclusion of more and varying clinical partners in the BioRef consortium, in principle both national and international, is easily feasible and advantageous. Once their data resources are aligned with the Swiss BioRef infrastructure, prospective partners can be granted access to the Swiss BioRef web interfaces in a very timely manner. An increase in total data volume will boost the chance to retrieve a sufficient number of patients for even very specific combinations of patient covariates, allowing to also run rather specialised data queries. In addition, expanding the set of included laboratory analytes and patient-specific parameters will allow for the definition of wider scientific questions when cross-sectional studies of RIs are defined. To better reflect the health status of patients, additional classification systems could be incorporated in the future. This could include e.g. the International Classification of Functioning, Disability and Health (ICF) which considers environmental factors relevant for the patient[48]. Such additional classifications could help to bridge normative and descriptive definitions of health and to introduce RIs to more clinical contexts.

### Sustainability

To secure long-term sustainability of the infrastructure, both costs for infrastructure operation and maintenance have been addressed from the start of the project. The simplified onboarding process for future partners to join affirms sustainability. As the Swiss BioRef tool is intended to support laboratory medicine, a continuation of the project within the framework of a professional clinical medicine society is intended. Similarly, the Swiss Society of Clinical Chemistry (SSCC) actively supports similar projects such as the <u>Hemolysis App</u>[49]. Third-party funding by the diagnostics industry, another potential beneficiary of precise RIs, is another option being followed up on. To maintain the MedCo codebase, a start-up founded at EPFL (<u>Tune Insight</u>) is available for support and customization of MedCo. Tune Insight has refined the system architecture of MedCo, promoting it now commercially as “TI4Health’’. A follow up-project “BioRef-MedCo” has been approved by SPHN for 2022, where participating partners of Insel, CHUV and Tune Insight collaborate to further industrialise the TI4Health system. With an industrialised TI4Health system instated, the Swiss BioRef infrastructure could be included as a subsidiary product of the BioMedIT network infrastructure in compliance with its ongoing IT-service optimization.

### Strength and limitations

The strength of the Swiss BioRef project lies in the successful establishment of a multi-cohort IT-resource for the estimation of precise reference intervals in laboratory medicine, the first of its kind in Switzerland and, to our knowledge, beyond. The project’s efforts have been manifold: Firstly, the various collaborators have been brought together into a multi-centre research consortium with a thoroughly defined common legal framework (CA) for multi-cohort data sharing. Secondly, common interoperable data formats and data encoding have been determined and implemented to secure the congruent fusion of the various local data sources into the unified Swiss BioRef infrastructure, both for a centralised database (Swiss BioRef Central) or an interlinked federated query system (Swiss BioRef MedCo). Lastly, the development of two capable web applications, with varying underlying data accessing architecture, allows physicians or clinical researchers to map the individual complexity of their patients into a substantiated statistical analysis to infer precise and highly relevant reference intervals independently of the data sharing policy of their hospital.

The differing data management systems and formats at the individual clinical data warehouses have been a limiting factor for smooth data provision: Significant effort has been required to harmonise the data contribution of all data providers and ensure interoperability prior to their availability to the Swiss BioRef infrastructure. As an example, implementing LOINC on a national level has advanced notably over the last years but still requires serious effort of providing high-quality metadata and quality control for laboratory analyses[33,34]. Another challenge will be the anticipation of future needs of an expanding and increasingly diverse user base. While it is possible to map a variety of differently complex analyses in the GUIs, extensive evaluations of the user experience and performance, including feedback from specialists in the field, will be required.

As of today, RIs for more than 40 blood test variables from clinical chemistry, haematology, point-of-care-testing, and coagulation can be estimated on-the-fly, where various patient factors (such as age, sex and a combination of ICD-10 defined diagnoses) and analytical factors (such as the GMDN type identifier or the GUDID unique identifier of analysers or test kits used in the clinical laboratories) can be incorporated into the analysis. The Swiss BioRef data set currently holds approximately 9 million measurements from 250’000 patients. The current approach in RI inference needs evaluations, as the inter-cohort transferability of semantic standards, “change tracking” in merged databases and biological variation of the blood test variables have not been thoroughly addressed. Given the modularity of both the BioRef consortium (future national and international partners can join the consortium with relative ease) and the applications (extendable for additional types of statistical analyses or variables) we see a bright future for personalised RIs in Switzerland - and beyond.

## Data Availability

Data is not available.

## Patient and public involvement

Patients and/or the public were not involved in the design, or conduct, or reporting, or dissemination plans of this research project.

## Patient consent for publication

Not applicable.

## Ethics waiver

KEK Bern, BASEC-Nr: Req-2020-00630.

## Provenance and peer review

Not commissioned; externally peer reviewed.

## Data availability statement

Primary data is not available. The reference interval analysis tools (working on aggregations of primary data) are accessible for Swiss researchers with a valid SWITCH edu-ID upon agreement on the Swiss BioRef end-user licence agreement (EULA) and registration with the Swiss BioRef team.

## Acknowledgments

The authors would like to thank their collaborators, in particular Simon Le Bail-Collet, their partners from the BioMedIT, DCC and SIB, Christos T. Nakas for continued support, and, most importantly, all patients who have given written consent for their data to be used in research.

